# Head-to-head performance comparison of self-collected nasal *versus* professional-collected nasopharyngeal swab for a WHO-listed SARS-CoV-2 antigen-detecting rapid diagnostic test

**DOI:** 10.1101/2021.03.17.21253076

**Authors:** Julian A.F. Klein, Lisa J. Krüger, Frank Tobian, Mary Gaeddert, Federica Lainati, Paul Schnitzler, Andreas K. Lindner, Olga Nikolai, B. Knorr, A. Welker, Margaretha de Vos, Jilian A. Sacks, Camille Escadafal, Claudia M. Denkinger, for the study team

**Affiliations:** Division of Clinical Tropical Medicine, Centre of Infectious Diseases, Heidelberg University Hospital, Germany; Department of Virology, Centre of Infectious Diseases, Heidelberg University Hospital, Germany; Charité – Universitätsmedizin Berlin, corporate member of Freie Universität Berlin, Humboldt-Universität zu Berlin, and Berlin Institute of Health, Institute of Tropical Medicine and International Health, Berlin, Germany; Local Health Authority of Heidelberg and Rhein-Neckar-Region, Germany; Foundation for Innovative New Diagnostics, Geneva, Switzerland

**Keywords:** SARS-CoV-2, COVID-19, nasal sampling, antigen-detecting rapid diagnostic test, self-sampling, head-to-head comparison

## Abstract

**Background:** In 2020, the World Health Organization (WHO) recommended two SARS-CoV-2 lateral flow antigen detecting rapid diagnostics tests (Ag-RDTs), both initially with nasopharyngeal (NP) sample collection. Independent head-to-head studies demonstrated for SARS-CoV-2 Ag-RDTs nasal sampling to be a comparable and reliable alternative for nasopharyngeal (NP) sampling.

**Methods:** We conducted a head-to-head comparison study of a supervised, self-collected nasal mid-turbinate (NMT) swab and a professional-collected NP swab, using the Panbio Ag-RDT (the second WHO-listed SARS-CoV-2 Ag-RDT, distributed by Abbott). We calculated positive and negative percent agreement and, compared to the reference standard reverse transcription polymerase chain reaction (RT-PCR), sensitivity and specificity for both sampling techniques.

**Results:** A SARS-CoV-2 infection could be diagnosed by RT-PCR in 45 of 290 participants (15.5%). Comparing the NMT and NP sampling the positive percent agreement of the Ag-RDT was 88.1% (37/42 PCR positives detected; CI 75.0% - 94.8%). The negative percent agreement was 98.8% (245/248; CI 96.5% - 99.6%). The overall sensitivity of Panbio with NMT sampling was 84.4% (38/45; CI 71.2% - 92.3%) and 88.9% (40/45; CI 76.5% - 95.5%) with NP sampling. Specificity was 99.2% (243/245; CI 97.1% - 99.8%) for both, NP and NMT sampling. The sensitivity of the Panbio test in participants with high viral load (> 7 log10 SARS-CoV-2 RNA copies/mL) was 96.3% (CI 81.7% - 99.8%) for both, NMT and NP sampling.

**Conclusion:** For the Panbio Ag-RDT supervised NMT self-sampling yields to results comparable to NP sampling. This suggests that nasal self-sampling could be used for scale-up population testing.

## To the Editor

The use of antigen-detecting rapid diagnostic tests (Ag-RDTs) for *SARS-CoV-2* has increased within the last months and has an important role in pandemic management. However, broader use and scale up is limited due to complex sampling methods. In 2020, the World Health Organization (WHO) recommended two lateral flow Ag-RDTs ((SD Biosensor, Inc. Gyeonggi-do, Korea, distributed by Roche, Germany, henceforth called Standard Q; and Abbott Panbio™ (Rapid Diagnostics, Jena, Germany; henceforth called PanBio)), both initially with nasopharyngeal (NP) sample collection [1,2]. Since then, independent head-to-head studies demonstrated that nasal sampling (including self-sampling) assessed against NP sampling leads to comparable performance using the *SARS-CoV-2* Ag-RDT SD STANDARD Q [3-5]. For Panbio, only one study to date assessed professional nasal mid-turbinate (NMT) sampling and showed 82.1% sensitivity and 99.1% specificity in comparison to reverse transcription polymerase chain reaction (RT-PCR). However, a head-to-head comparison with NP sampling has not been performed to date [6].

We conducted a manufacturer-independent prospective study directly comparing the diagnostic accuracy of Panbio performed with a supervised, self-collected NMT swab versus a professionally collected NP swab. For the two Ag-RDT sampling techniques positive and negative percent agreements (PPA, NPA) were calculated. Sensitivity and specificity were assessed and compared against the reference standard RT-PCR.

The ethical review committee at Heidelberg University Hospital approved the study protocol (registration number S-180/2020). Enrollment and testing took place in Heidelberg (Germany) between December 15^ths^ 2020 and January 19^ths^ 2021 in a *SARS-CoV-2* drive-in testing centre, led by the local health authority. We included adults with symptoms suggestive for a *SARS-CoV-2* infection or a recent high-risk contact with a confirmed *SARS-CoV-2* case. After written informed consent, each participant was instructed to self-collect a NMT swab for the Ag-RDT under supervision using a non-flocked swab (Jiangsu Changfeng Medical Industry Co., Ltd., Jiangsu, China), provided by Abbott in the research use only Panbio kit for nasal swab testing. The instructions were verbal and picture-guided following the manufacturer’s instructions for use. In a second step, a health worker collected a NP swab (using IMPROSWAB®, Guangzhou Improve Medical Instruments Co., Ltd., Guangzhou, China), for RT-PCR testing in one nostril. Finally, a second NP swab for Ag-RDT testing was collected from the patient using a nylon-flocked specimen (NFS-SWAB Applicator™, Noble Bioscienes Inc., Gyeonggi-do, Korea), provided with the commercial Abbott (nasopharyngeal) test kit. The PanBio was conducted on-site by trained study personnel following the manufacturer’s instruction for use for each kit [7]. Two study staff read out the Ag-RDT results, each of them blinded to the interpretation of the other.

For RT-PCR testing the Tib Molbiol® (Berlin, Germany) assay was used. Viral load (VL) values were calculated based on a calibration curve and the assay specific cycle threshold (Ct)-value [8]. Leftover samples of the NMT and NP swabs resuspended in Ag-RDT buffer were stored at −20 degrees Celsius. Samples that were identified to be false-positive in comparison to RT-PCR on one or both Ag-RDTs were retested with RT-PCR from the remnant Ag-RDT buffer.

We screened a total of 369 eligible individuals of whom 292 (79.1%) gave written consent. After exclusion of two participants (one with invalid RT-PCR result and one with lost written informed consent), 290 participants were included in the analysis (study flow detailed in Supplementary Material (B)). Our study population had an average age of 42.7 years (standard deviation (SD) 14.6), 33.8% (98/290) had comorbidities and 52.4% (152/290) were female. In total, 45.9% (133/290) were symptomatic on the day of testing with a mean duration of symptoms of 3.8 days (SD 5.4). *SARS-CoV-2* infection was detected by RT-PCR in 15.5% (45/290) of the study population (Table 1), with eight infections being among asymptomatic participants. One invalid Ag-RDT was registered on NP samples, which was valid upon repeat.

**TABLE 1.** Ag-RDT results with a supervised self-collected nasal mid-turbinate (NMT) swab and professional-collected nasopharyngeal (NP) swab in RT-PCR positive patients.

| Ag-RDT (NMT swab)<br>self-collected | Ag-RDT (NP swab)<br>prof.-collected | RT-PCR<br>(NP swab)<br>professionell collected |  | Symptom<br>duration<br>(days) |
| --- | --- | --- | --- | --- |
|  |  | Ct-value* | Viral load<br>(log<sub>10</sub><br>SARS-CoV-2 RNA<br>copies/ml) |  |
| positive | positive | 12.7 | 10.0 | 2 |
| positive | positive | 12.9 | 9.9 | 4 |
| positive | positive | 13.1 | 9.9 | 3 |
| positive | positive | 16.1 | 9.0 | 3 |
| positive | positive | 16.4 | 8.9 | 3 |
| positive | positive | 16.5 | 8.9 | 2 |
| positive | positive | 16.5 | 8.9 | 1 |
| positive | positive | 16.6 | 8.9 | 1 |
| positive | positive | 16.7 | 8.8 | 1 |
| positive | positive | 17.8 | 8.5 | 0 |
| positive | positive | 17.9 | 8.5 | 1 |
| positive | positive | 18.8 | 8.2 | 2 |
| positive | positive | 18.8 | 8.2 | 7 |
| positive | negative | 18.9 | 8.2 | 1 |
| positive | positive | 19.5 | 8.0 | 5 |
| positive | positive | 19.7 | 7.9 | 1 |
| positive | positive | 19.9 | 7.9 | 4 |
| positive | positive | 19.9 | 7.9 | asymptomatic<sup>#</sup> |
| positive | positive | 20.1 | 7.8 | asymptomatic<sup>#</sup> |
| positive | positive | 20.2 | 7.8 | 10 |
| positive | positive | 21.2 | 7.5 | 2 |
| positive | positive | 21.4 | 7.4 | asymptomatic<sup>#</sup> |
| negative | positive | 22.1 | 7.2 | 1 |
| positive | positive | 22.5 | 7.1 | 5 |
| positive | positive | 22.5 | 7.1 | 2 |
| positive | positive | 22.6 | 7.1 | 6 |
| positive | positive | 22.8 | 7.0 | 5 |
| positive | positive | 23.1 | 6.9 | asymptomatic<sup>#</sup> |
| positive | positive | 23.1 | 6.9 | 1 |
| positive | positive | 23.6 | 6.8 | 5 |
| positive | positive | 23.8 | 6.7 | 2 |
| positive | positive | 25.7 | 6.2 | 2 |
| positive | positive | 25.9 | 6.1 | 7 |
| positive | positive | 26.0 | 6.1 | 6 |
| positive | positive | 26.3 | 6.0 | asymptomatic<sup>#</sup> |
| positive | positive | 26.7 | 5.9 | 1 |
| negative | negative<sup>1</sup> | 26.7 | 5.9 | 1 |
| positive | positive | 27.7 | 5.6 | 2 |
| negative | positive | 29.7 | 5.0 | asymptomatic<sup>#</sup> |
| negative | negative | 30.6 | 4.7 | asymptomatic<sup>#</sup> |
| positive | positive | 31.2 | 4.5 | 1 |
| positive | negative | 31.2 | 4.5 | 1 |
| negative | positive | 32.7 | 4.1 | n.a. |
| negative | positive | 33.8 | 3.8 | asymptomatic<sup>#</sup> |
| negative | negative | 34.5 | 3.6 | 10 |
| Sensitivity<br>84.4% (38/45;<br>CI 71.2% – 92.3%) | Sensitivity<br>88.9% (40/45;<br>CI 76.5% – 95.5%) |  |  |  |
| Positive Percent Agreement<br>88.1% (CI 75.0% – 94.8%)** |  |  |  |  |

*\*Assay: TibMolBiol*
*\*\*including one false-positive on NMT and NP and one on NP*
*<sup>#</sup>on the day of testing*
*<sup>1</sup>oropharyngeal swab due to contraindications of NP Sampling*
**Abbreviations:** Ag-RDT: antigen-detecting rapid diagnostic test; NMT: nasal mid-turbinate;
NP: nasopharyngeal; Ct: cycle threshold; RT-PCR: reverse transcription-polymerase chain
reaction; n.a.: not available. CI: confidence interval

The overall sensitivity of Panbio with NP sampling was 88.9% (40/45; 95% confidence interval (CI) 76.5% - 95.5%) and 84.4% (38/45; CI 71.2% - 92.3%) with NMT sampling. Four infections were identified by NP Ag-RDT sampling, which were negative in NMT sampling of which two had a low VL (VL <4.9 log_10_ *SARS-CoV-2* RNA copies/ml) and two were asymptomatic (Table 1). Two participants had a positive NMT result, not detected via NP Ag-RDT, of which one had a low VL (VL <4.9 log_10_ *SARS-CoV-2* RNA copies/ml). Specificity was 99.2% (243/245; CI 97.1% - 99.8%) for both, NP and NMT sampling. Considering only RT-PCR positive participants with high VL (> 7 log_10_ *SARS-CoV-2* RNA copies/mL), the sensitivity of the Panbio test was 96.3% (CI 81.7% - 99.8%) for both NMT and NP sampling. Excluding nine participants with oropharyngeal sampling instead of NP (due to contraindications of NP sampling) increased sensitivity only marginally (40/44; 90.9% (CI 78.8% – 96.4%)). Detailed results by symptoms and sub-group analyses are available in the Supplement (C&D). The positive percent agreement of the Ag-RDT was 88.1% (37/42 PCR positives detected; CI 75.0% - 94.8%) including one false-positive by both NMT and NP, and one false-positive by NP only. The negative percent agreement was 98.8% (245/248; CI 96.5% - 99.6%). Inter-rater reliability for the interpretation of the Ag-RDTs was perfect with a kappa of 1.0. Participants reported NMT sampling to be better tolerated than NP sampling.

When performing RT-PCR from the remnant buffer/sample-mixture, SARS CoV-2 was identified in both NMT and NP samples from the same participant with a false-positive Ag-RDT result. This suggests the Ag-RDT result being in fact true-positive with a sampling error likely having occurred for the RT-PCR from NP sample. For two other false-positives, one each on NMT and NP, no virus was identified in the buffer solution. Among three false-negative NMT samples one buffer was positive with low VL (4.38 log_10_ *SARS-CoV-2* RNA copies/ml; Supplementary Material (F), suggesting that the VL was below the limit of detection of the Ag-RDT.

Our study has several strengths. Study methods were rigorous and included standardized sampling and two independent blinded readers. The study population is representative, judging from the similar sensitivity of the Panbio test with NP sampling observed in our study in comparison to two large validation studies [9,10]. All samples for routine RT-PCR were tested via the same RT-PCR assay (Table 1). The RT-PCR on the leftover buffer solution of Ag-RDT allowed us to perform further discrepant analysis.

A limitation of the study is that it was performed in a single centre. The preselection of participants invited to come for testing was done according to national guidelines. We did not record deviations from the recommended NMT procedure, however as the sampling was done under proactive supervision, no major deviations were observed. Readers were not blinded to the sampling method while interpreting the test results, but weak positive results are rarely observed with the Panbio test, thus this limitation is unlikely to result in a difference in result interpretation. In the discrepant analysis, we did not perform RT-PCR of all Ag-RDT buffer solutions thus introducing a possible bias.

Our study suggests that supervised NMT self-sampling leads to results comparable to NP sampling for the PanBio Ag-RDT. A possible reduction in VL present in the nasal region compared to the nasopharyngeal region may be counterbalanced by the ease-of-sampling. Results of nasal sampling could potentially be further improved, if flocked swabs were used [11]. Standardized easy self-sampling methods are highly desirable, as they could increase throughput and require fewer medical personnel, which is often a bottle neck for scaling of antigen testing.

## Data Availability

All raw data and analysis code are available upon a request to the corresponding author.

## Acknowledgements

Angelika Sandritter, Andrea Sieber, Alexander Syring, Zoe Solomon, Emilija Mitreska, Sabrina Eisenmann, Andrea Fuhs, Kholoud Assaad, Salome Steinke.

