## Supplementary Material for "Head-to-head performance comparison of self-collected nasal *versus* professional-collected nasopharyngeal swab for a WHO-listed SARS-CoV-2 antigen-detecting rapid diagnostic test"

### (A) Table 1: Study Team

| | |
|---|---|
| Public Health Authority, Rhein-Neckar-Region, Heidelberg, Germany | Dr. K. Assaad, |
| | Dr. A. Fuhs |
| | Dr. C. Harter |
| | C. Schulze |
| | G. Schmitt |
| Division of Clinical Tropical Medicine, Heidelberg University Hospital, Heidelberg Germany | Loai Abutaima |
| | Rico Müller |
| | Martina Fink |
| | Mathilde Fougereau |
| | Maximilian Schirmer |
| | Annika Small |
| | Matthias Meinlschmidt |
| | Valerie Dürr |
| | Alina Schuckert |
| | Ann-Kathrin Backes |
| | Salome Steinke |
| | Henrik Ellinghaus |
| | Magdalena Mikula |
| | Nele Schäfer |

### (B) Figure 1: Study Flow

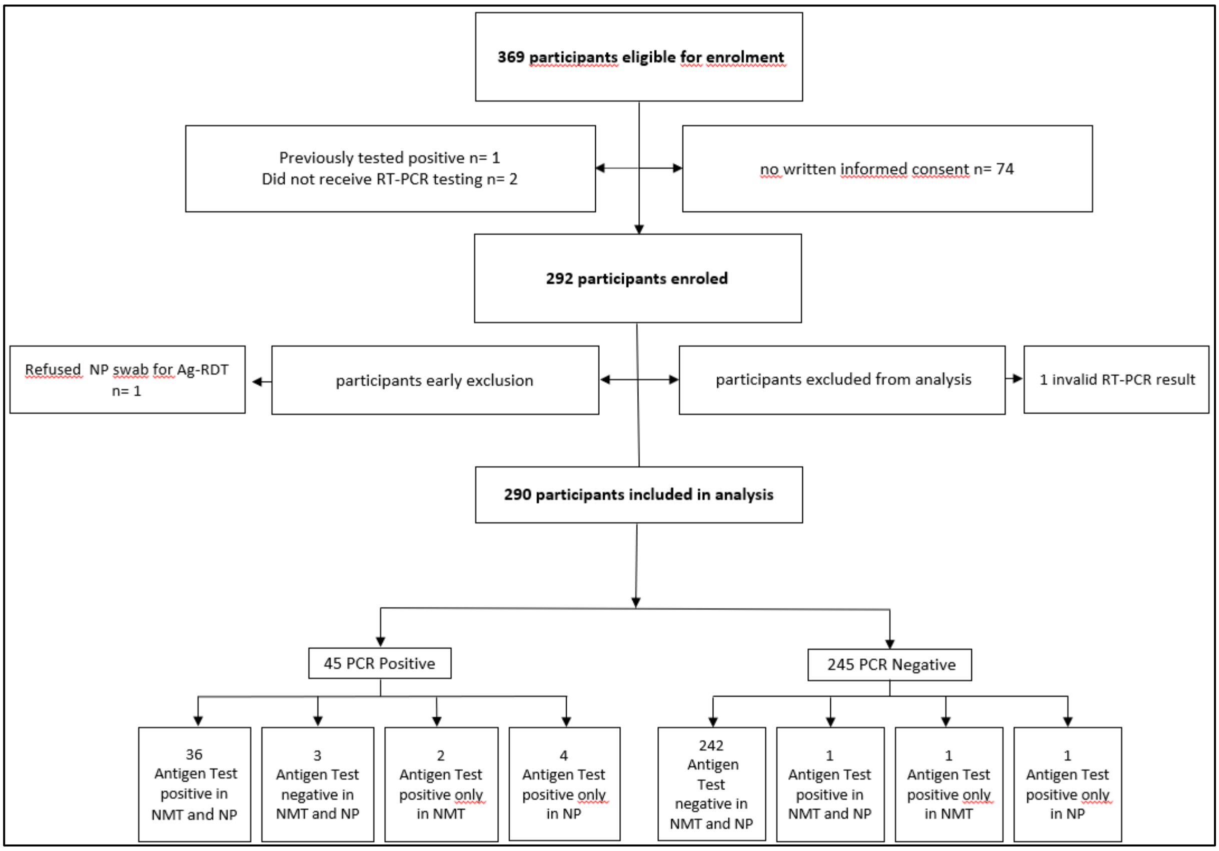

### (C) Table 2: Detailed list of symptoms for all PCR positive participants

| CT value: E-Gene | Viral load ($log_{10}$RNA copies/mL) | Result Ag-RDT NMT swab | Result Ag-RDT NP swab | Increased temperature/ fever | Cough | Productive Cough | Sore throat | Dyspnoe | Body aches/ muscle pain | Fatigue | Headache | Runny nose | Chest pain | Diarrhea | Nausea / Vomitus | Loss of taste/ smell | Other symptoms |
|---|---|---|---|---|---|---|---|---|---|---|---|---|---|---|---|---|---|
| 13.1 | 9.9 | positive | positive | No | Yes | No | No | No | Yes | Yes | Yes | No | No | No | No | No | Yes |
| 25.7 | 6.2 | positive | positive | No | Yes | No | No | No | Yes | Yes | Yes | No | No | Yes | No | No | No |
| 22.6 | 7.1 | positive | positive | Yes | Yes | No | Yes | No | Yes | Yes | Yes | No | Yes | No | No | Yes | No |
| 31.2 | 4.5 | positive | negative | No | No | No | Yes | No | Yes | Yes | Yes | No | No | No | No | No | No |
| 26.0 | 6.1 | positive | positive | No | No | No | No | No | No | No | Yes | Yes | No | No | No | Yes | No |
| 22.1 | 7.2 | negative | positive | Yes | Yes | No | Yes | No | Yes | Yes | Yes | No | No | Yes | Yes | No | |
| 23.1 | 6.9 | positive | positive | | | | | | | | | | | | | | |
| 26.7 | 5.9 | positive | positive | No | No | No | No | No | No | No | No | No | No | No | No | Yes | No |
| 16.1 | 9.0 | positive | positive | No | Yes | Yes | Yes | No | Yes | Yes | No | Yes | No | No | No | No | No |
| 16.5 | 8.9 | positive | positive | Yes | Yes | No | Yes | | No | Yes | Yes | No | No | No | No | No | No |
| 19.7 | 7.9 | positive | positive | No | No | No | Yes | No | No | Yes | Yes | No | No | No | No | No | No |
| 26.7 | 5.9 | negative | negative | No | No | No | No | No | No | No | Yes | No | No | No | No | No | No |
| 16.6 | 8.9 | positive | positive | Yes | No | No | No | No | No | No | Yes | No | No | No | No | | No |
| 18.9 | 8.2 | positive | negative | No | No | No | No | No | No | No | No | Yes | No | No | No | No | Yes |
| 23.8 | 6.7 | positive | positive | No | No | No | No | No | Yes | Yes | No | Yes | | No | No | Yes | No |
| 16.7 | 8.8 | positive | positive | No | Yes | No | Yes | No | Yes | Yes | Yes | No | Yes | No | No | No | No |
| 17.8 | 8.5 | positive | positive | Yes | No | No | No | No | Yes | No | No | No | No | No | No | No | No |
| 20.1 | 7.8 | positive | positive | | | | | | | | | | | | | | |
| 16.4 | 8.9 | positive | positive | No | Yes | | Yes | No | No | Yes | Yes | No | No | No | No | No | No |
| 30.6 | 4.7 | negative | negative | | | | | | | | | | | | | | |
| 17.9 | 8.5 | positive | positive | Yes | No | No | No | No | Yes | Yes | Yes | No | No | No | No | No | No |
| 21.2 | 7.5 | positive | positive | Yes | No | Yes | Yes | No | Yes | Yes | Yes | No | Yes | No | No | No | Yes |
| 18.8 | 8.2 | positive | positive | Yes | No | No | Yes | No | Yes | Yes | Yes | Yes | No | No | No | No | No |
| 27.7 | 5.6 | positive | positive | Yes | Yes | Yes | No | No | Yes | Yes | Yes | No | No | No | No | Yes | No |
| 12.7 | 10.0 | positive | positive | No | Yes | Yes | No | No | No | Yes | Yes | Yes | No | No | No | No | No |
| 16.5 | 8.9 | positive | positive | No | Yes | Yes | No | No | Yes | No | Yes | Yes | No | Yes | No | No | No |
| 33.8 | 3.8 | negative | positive | | | | | | | | | | | | | | |
| 22.5 | 7.1 | positive | positive | No | No | No | No | No | Yes | No | Yes | No | No | No | No | Yes | No |

| CT value: E-Gene | Viral load ($log_{10}$RNA copies/mL) | Result Ag-RDT NMT swab | Result Ag-RDT NP swab | Increased temperature/ fever | Cough | Productive Cough | Sore throat | Dyspnoe | Body aches/ muscle pain | Fatigue | Headache | Runny nose | Chest pain | Diarrhea | Nausea / Vomitus | Loss of taste/ smell | Other symptoms |
|---|---|---|---|---|---|---|---|---|---|---|---|---|---|---|---|---|---|
| 12.9 | 9.9 | positive | positive | Yes | Yes | Yes | Yes | No | No | Yes | No | Yes | No | No | No | No | No |
| 25.9 | 6.1 | positive | positive | Yes | Yes | Yes | No | No | Yes | Yes | Yes | Yes | No | No | No | No | No |
| 18.8 | 8.2 | positive | positive | Yes | No | No | No | No | Yes | Yes | Yes | No | No | No | No | Yes | Yes |
| 23.6 | 6.8 | positive | positive | | Yes | No | Yes | No | Yes | Yes | Yes | Yes | No | No | No | Yes | |
| 34.5 | 3.6 | negative | negative | No | Yes | No | No | No | No | No | No | Yes | No | No | No | No | No |
| 19.9 | 7.9 | positive | positive | No | Yes | No | Yes | No | No | Yes | Yes | No | No | Yes | No | Yes | No |
| 23.1 | 6.9 | positive | positive | No | Yes | Yes | Yes | No | Yes | Yes | Yes | Yes | No | No | Yes | No | No |
| 31.2 | 4.5 | positive | positive | No | No | No | No | No | No | Yes | Yes | Yes | No | Yes | No | Yes | No |
| 21.4 | 7.4 | positive | positive | | | | | | | | | | | | | | |
| 29.7 | 5.0 | negative | positive | | | | | | | | | | | | | | |
| 22.8 | 7.0 | positive | positive | No | Yes | No | No | No | Yes | Yes | Yes | No | Yes | No | No | Yes | No |
| 22.5 | 7.1 | positive | positive | Yes | Yes | Yes | No | Yes | No | Yes | Yes | No | No | No | No | No | No |
| 19.9 | 7.9 | positive | positive | | | | | | | | | | | | | | |
| 26.3 | 6.0 | positive | positive | | | | | | | | | | | | | | |
| 32.7 | 4.1 | negative | positive | Yes | Yes | No | Yes | Yes | Yes | Yes | Yes | Yes | Yes | No | No | Yes | No |
| 20.2 | 7.8 | positive | positive | Yes | Yes | No | No | No | Yes | Yes | Yes | Yes | No | No | No | Yes | No |
| 19.5 | 8.0 | positive | positive | No | Yes | No | No | No | Yes | No | Yes | Yes | No | No | No | No | No |

### (D) Table 3: Sensitivity and Specificity overall and by subgroups

| | Sampling technique | Sensitivity (%; 95% CI) | Specificity (%; 95% CI) | Positive Percent Agreement | Negative Percent Agreement |
|---|---|---|---|---|---|
| **Overall** | NP | 40* / 45 (88.9%; 76.5% - 95.5%) | 243 / 245 (99.2%; 97.1% - 99.8%) | 37* / 42** (88.1%; 75.0% - 94.8%) | 245 / 248 (98.8%; 96.5% - 99.6%) |
| | NMT | 38 / 45 (84.4%; 71.2% - 92.3%) | 243 / 245 (99.2%; 97.1% - 99.8%) | | |
| **Viral load ≥7 log10** *SARS-CoV-2 RNA copies/ml* | NP | 26 / 27 (96.3%; 81.7% - 99.8%) | n.a. | 25 / 26 (96.2%; 81.1% - 99.8%) | n.a. |
| | NMT | 26 / 27 (96.3%; 81.7% - 99.8%) | n.a. | | |
| **Viral load <7 log10** *SARS-CoV-2 RNA copies/ml* | NP | 14 / 18 (77.8%; 54.8% - 90.1%) | n.a. | 11 / 14 (78.6%; 52.4% - 92.4%) | n.a. |
| | NMT | 12 / 18 (66.7%; 43.7% - 83.7%) | n.a. | | |
| **symptomatic** | NP | 33/37 (89.2%, 75.3% - 95.7%) | 94/96 (97.9%, 81.4% - 99.4%) | 33 / 35 (91.4%, 77.6% - 97.0%) | 96 / 98 (98.0%, 92.8% - 99.4%) |
| | NMT | 33/37 (89.2%, 75.3% - 95.7%) | 95/96 (99.0%, 94.3% - 99.9%) | | |
| **asymptomatic** | NP | 7/8 (87.5%, 52.9% - 99.4%) | 147/147 (100%, 97.5 - 100%) | 5 / 7 (71.4%, 35.9% - 91.8%) | 147 / 148 (99.3%, 96.3% - 100%) |
| | NMT | 5/8 (62.5%, 30.6% - 86.3%) | 146/147 (99.3%, 96.2% - 100%) | | |

Abbreviations: n.a.: not applicable; NP: nasopharyngeal; NMT: nasal mid-turbinate; CI: confidence interval

**including one false-positive in NMT and NP

**including two false-positive in NP

### (E) Table 4: Ag-RDT – RT-PCR discrepant analysis: Buffer solution RT-PCR-results of Ag-RDT false-positive and Ag-RDT false-negative retained samples

**Ag-RDT false-positives**

| Ag-RDT NMT swab | Ag-RDT NP swab | RT-PCR | Assay | RT-PCR buffer NMT sample | RT-PCR buffer NP sample | Sampling comment | Probe aspect | Symptom duration (days) |
|---|---|---|---|---|---|---|---|---|
| positive | negative | negative | TibMolBiol | negative | negative | none | NMT: mucous NP: clear | asymptomatic |
| positive | positive | negative | TibMolBiol | positive (Ct 27.3; VL* 6.3) | positive (Ct 31.8; VL* 5.0 | none | NMT: clear NP: mucous | 3 |
| negative | positive | negative | TibMolBiol | negative | negative | none | NMT: clear NP: mucous | 6 |

**Ag-RDT false-negatives**

| Ag-RDT NMT swab | Ag-RDT NP swab | Ct-value | Viral load* | Assay | RT-PCR buffer NMT sample | RT-PCR buffer NP sample | Sampling comment | Probe aspect | Symptom duration (days) |
|---|---|---|---|---|---|---|---|---|---|
| negative | positive | 22.1 | 7.2 | TibMolBiol | sample got lost | not tested | none | NMT: clear NP:clear | 1 |
| negative | positive | 29.7 | 5.0 | TibMolBiol | negative | not tested | none | NMT: mucous NP:clear | asymptomatic |
| negative | positive | 32.7 | 4.1 | TibMolBiol | positive (Ct 33.7; VL* 4.4) | not tested | none | NMT: mucous NP: mucous,bloody spots | n.a. |
| negative | positive | 33.8 | 3.8 | TibMolBiol | negative | not tested | none | NMT: clear NP: mucous | asymptomatic |
| positive | negative | 18.9 | 8.2 | TibMolBiol | not tested | negative | none | NMT: clear NP:clear | 1 |
| positive | negative | 31.2 | 4.5 | TibMolBiol | not tested | sample got lost | none | NMT: clear NP:clear | 1 |

Ct: cycle threshold; Ag-RDT: antigen-detecting rapid diagnostic test; NMT: nasal mid-turbinate; NP: nasopharyngeal. n.a.: not available; *$\log_{10}$ *SARS-CoV-2* RNA copies/ml
